# Prospective, Randomized, Parallel-Group, Open-Label Study to Evaluate the Efficacy and Safety of IMU-838, in Combination with Oseltamivir, in Adults with Coronavirus-19 – The IONIC Trial Protocol

**DOI:** 10.1101/2021.07.13.21258757

**Authors:** Kavi Sharma, Lisa Berry, Evangelos Vryonis, Asad Ali, Beatriz Lara, Angela Noufaily, Nick Parsons, Christopher James Bradley, Becky Haley, Tabuso Maria, Ramesh Arasaradnam

## Abstract

**Background:** Globally there is a scarcity of effective treatments for SARS-CoV-2 infections (causing COVID 19). Repurposing existing medications may offer the best hope for treating COVID 19 patients to curb the pandemic. IMU-838 is a dihydroorotate dehydrogenase (DHODH) inhibitor, which is an effective mechanism for antiviral effects against respiratory viruses. When used synergistically with Oseltamivir, therapeutic effects have been observed against influenza and SARS-CoV-2 in rodents.^(13)^ The IONIC trial is a randomized control trial that will investigate whether time to clinical improvement in COVID 19 patients is improved following a 14 day course of IMU-838 + Oseltamivir versus Oseltamivir alone.

**Methods:** IONIC trial is an open label study in which participants will be randomised 1:1 in two parallel arms; the intervention arm (IMU-838 + Oseltamivir) and control arm (Oseltamivir only). The primary outcome is time-to-clinical improvement; defined as the time from randomisation to: a 2-point improvement on WHO ordinal scale; discharge from hospital, or death (whichever occurs first). The study is sponsored by UHCW NHS Trust and funded by LifeArc.

**Discussion:** The IONIC Protocol describes an overarching trial design to provide reliable evidence on the efficacy of IMU-838 (vidofludimus calcium) when delivered in combination with an antiviral therapy (Oseltamivir) [*IONIC Intervention*] for confirmed or suspected COVID-19 infection in adult patients receiving usual standard of care.

**Trial Registration:** The trial was registered with EudraCT (2020-001805-21) on 09.04.2020 and ISRCTN on 23.09.2020 (<u>ISRCTN53038326</u>) and Clinicaltrials.gov on 17.08.2020 (NCT04516915)

**Strengths and Limitations:** This study is the first to recruit participants in the trial exploring the effectiveness of IMU-838 in COVID-19. In addition, we believe it is the only trial exploring the effectiveness of IMU-838 in combination with Oseltamivir (Tamiflu) in patients with moderate to severe COVID-19. However, to make the trial design flexible due to the on-going pandemic the trial is un-blinded.

## 1. Background

### 1.1 Background and Justification

The World Health Organization (WHO) declared severe acute respiratory syndrome coronavirus (SARS-CoV-2) infections (causing coronavirus disease 2019 [COVID-19]) a pandemic on March 11, 2020. Main clinical symptoms include fever, cough, myalgia or fatigue, expectoration, and dyspnoea^(1).^ While a majority of patients do not experience severe symptoms, one early meta-analysis found that approximately 18% of cases were severe^(2)^ with a fatality rates estimated to be ∼4-7% at this time^(2,3)^. A more recent meta-analysis suggests fatality rates of COVID 19 are around 0.68% (Meyerowitz-Katz & Merone, 2020).

At the time of study conception, there were no known treatments for COVID-19. Whilst the anticipated scale of the epidemic is such that hospitals, and particularly intensive care facilities, may be massively overstretched. As described by a few models of pandemic spread, up to 50% of an adult population may fall sick over a period of 8-12 weeks without intervention, of whom around 10% may require hospitalisation. This figure could imply nearly 2 million hospital admissions in the UK alone. Considering this scenario, therapies which may only have a moderate impact on survival or on hospital resources should be worth investigating.

The IONIC Protocol describes an overarching trial design to provide reliable evidence on the efficacy of IMU-838 (vidofludimus calcium) when delivered in combination with an antiviral therapy (Oseltamivir) [*IONIC Intervention*] for confirmed or suspected COVID-19 infection in hospitalised adult patients receiving usual standard of care.

### 1.2 Choice of Intervention

#### IMU-838

Vidofludimus free acid (SC12267) was previously developed by 4SC AG using capsules or tablets containing amorphous vidofludimus (4SC-101). Immunic AG acquired all rights and data of SC12267 and have developed a new pharmaceutical form containing the calcium salt of vidofludimus (INNM: vidofludimus calcium) in a new pharmaceutical formulation (tablets containing a specific polymorph).

### 1.3 Safety of IMU-838

To date, 351 individuals have been exposed to vidofludimus (not including the ongoing and still blinded Phase 2 trial in RRMS). Of these 351 subjects, 299 were dosed with 4SC-101 and 52 with IMU-838.

The safety analysis of all exposed subjects provided the following findings: No deaths, no serious adverse events during Phase 1 with IMU-838.

The most frequent adverse events for IMU-838 during Phase 1 were: headaches, flatulence, common cold symptoms, and positive urine dipstick for haemoglobin. Importantly, vidofludimus (free acid) at a daily dose of 35 mg showed no increase of adverse reactions compared with placebo, and no increased infection rate.

### 1.4 IMU-838 and COVID-19 (SARS-CoV-2)

IMU-838 selectively inhibits pyrimidine synthesis via inhibition of DHODH, which may be promising approach to treat COVID-19. Inhibition of de novo pyrimidine biosynthesis is a well-recognized mechanism of action associated with antiviral effects against respiratory viruses. ^(5–11)^ The presumptive explanation is attributed to the direct depletion of host nucleosides necessary for replication of the viral genome; however, secondary activation of the innate immune response has also been described as a relevant downstream mechanism. ^(6,11,12)^ Pyrimidine depletion is primarily achieved by blocking DHODH, an enzyme involved in the rate-limiting step of pyrimidine biosynthesis. Therefore, DHODH inhibition ameliorates and blocks the viruses’ ability to “hijack” the human host cells mechanism of RNA production as a means to virus replication. Further detail of in vitro and in vivo trials is shown in Appendix 1.

### 1.5 IMU-838 and Oseltamivir

The data described by Xiong et al. ^(13)^ described the synergistic response between a DHODH inhibitor (where IMU-838 is one such example) and Oseltamivir in Influenza infected mice. Specific inhibition of SARS-COV-2 was shown with DHODH inhibitors alone but not with Oseltamivir. In particular, IMU-838 was shown to have a clear activity against SARS-CoV-2 in cellular assays at mid-range single-digit micromolar range. This activity is well below the plasma concentrations of IMU-838 with the dosing regimen proposed in this trial (see figure 1).

**Figure 1.**
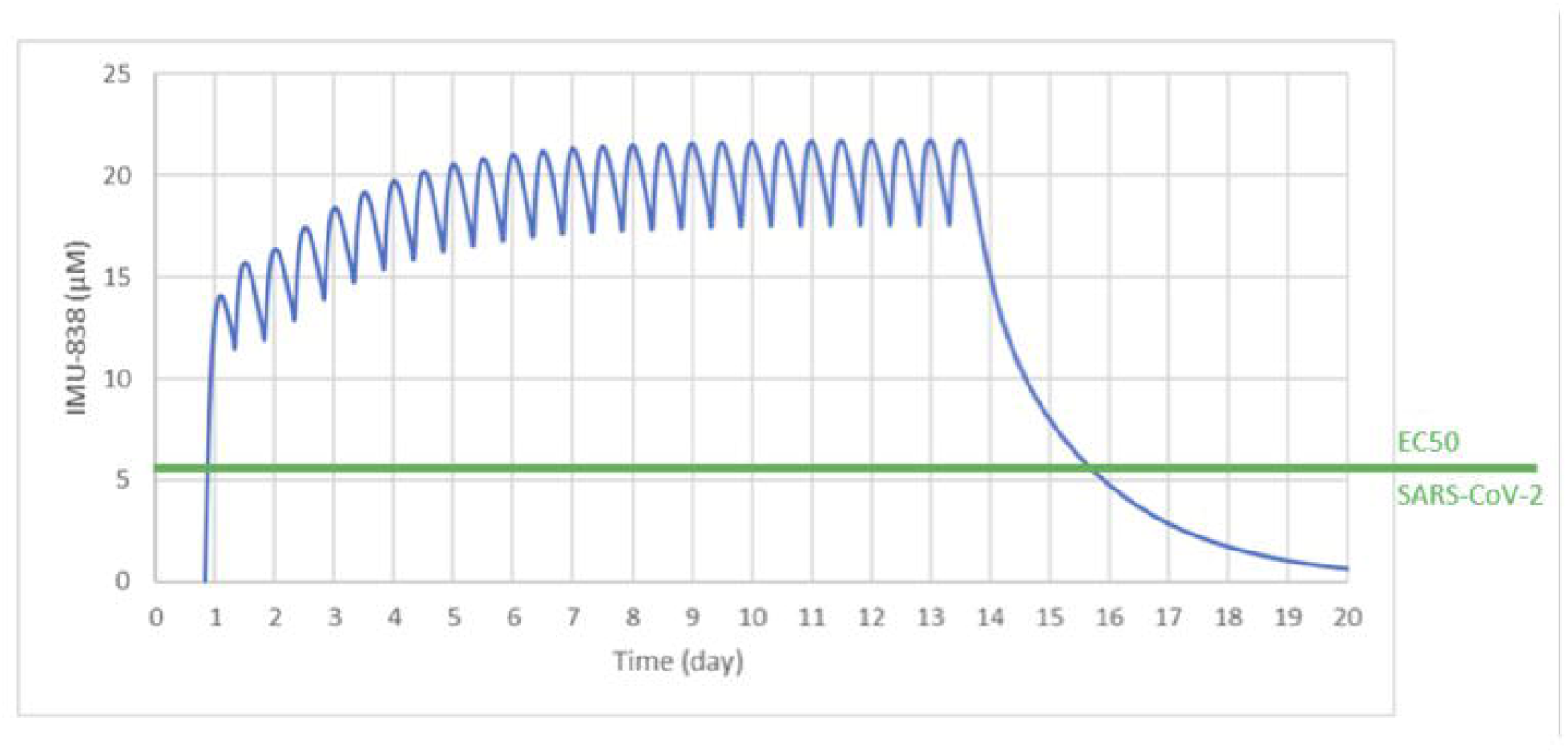
Pharmakokinetic profile for 22.5mg BD of IMU-838 over a 14 day treatment period.

While there is no data at present demonstrating direct activity of Oseltamivir against SARS-COV-2; the IONIC trial is investigating the combination effect of IMU-838 and Oseltamivir and in this regard, the Oseltamivir only arm represents the control arm. The trial is not about investigating the effect of Oseltamivir on SARS-COV-2.

An important consideration is that Influenza is a recurring infection with reports of co-infection with SARS-COV-2 ^(14, 15)^. Hence it would seem prudent to protect patients in both arms including the ‘control arm’ of this possibility. In fact Ding et al.^(14)^ reported use of Oseltamivir in addition to standard care in patients with SARS-COV-2 co-infected with Influenza. Of particular note, Oseltamivir is usually given early in a viral infection. Xiong et al. ^(13)^ data shows that IMU-838 re-sensitizes Oseltamivir to also be effective in the later stages of virus infection which is very important for this proposed trial. We can extrapolate its effects to that of SARS-COV-2 based on the assumption that another drug (Favipiravir) of the same class as Oseltamivir has shown a clinical relevant effect in COVID-19 patients in a trial in China ^(16)^. Moreover, the recent report by Costanzo et al. ^(16)^ demonstrates the synergistic effect of Oseltamivir (in this case when combined with Lopinavir/Ritonavir) in the treatment of COVID-19 lending support to our rationale that it is the synergistic effect of Oseltamivir with either an antiviral or DHODH inhibitor that seems effective. A further consideration: we also know that gastrointestinal symptoms can affect up to 60% ^(17)^ of those with COVID and a systematic review of Oseltamivir (in Influenza) ^(18)^ has shown reduction in the proportion with diarrhoea. Hence we perceive this to be an added therapeutic benefit.

If this fixed combination therapy (IMU-838 and Oseltamivir) is proven to be effective against COVID-19, it would also offer a more cost-effective treatment option in the long term compared to other anti-virals as Oseltamivir is cheap and is easily available. We did explore other anti-viral remedies such as Remdesivir and Favipiravir but these are not available in UK or Europe at the time of study conception. Hence the practicalities of having an available drug in stock in the UK have been given considerable weighting when designing this project.

In an ideal scenario, we would repeat the experiments of Xiong et al. against SARS-COV-2 using Oseltamivir but the urgency of this pandemic precludes this hence we have adopted a practical approach based on the best available evidence. It is for the above reasons we have chosen to add Oseltamivir within the control arm.

## 2. Methodology

### 2.1. Trial Procedures

The IONIC trial is an interventional, randomised, parallel-group, open-label, Phase IIb trial to assess the efficacy and safety of an oral dose of IMU-838 (22.5 mg twice daily [45 mg/day]) plus Oseltamivir (75mg twice daily [150mg/day]) (IONIC Intervention) in comparison with Oseltamivir alone (75mg twice daily) for 14 days in hospitalised patients with COVID-19. Figure 2 illustrates the design of the trial.

**Figure 2:**
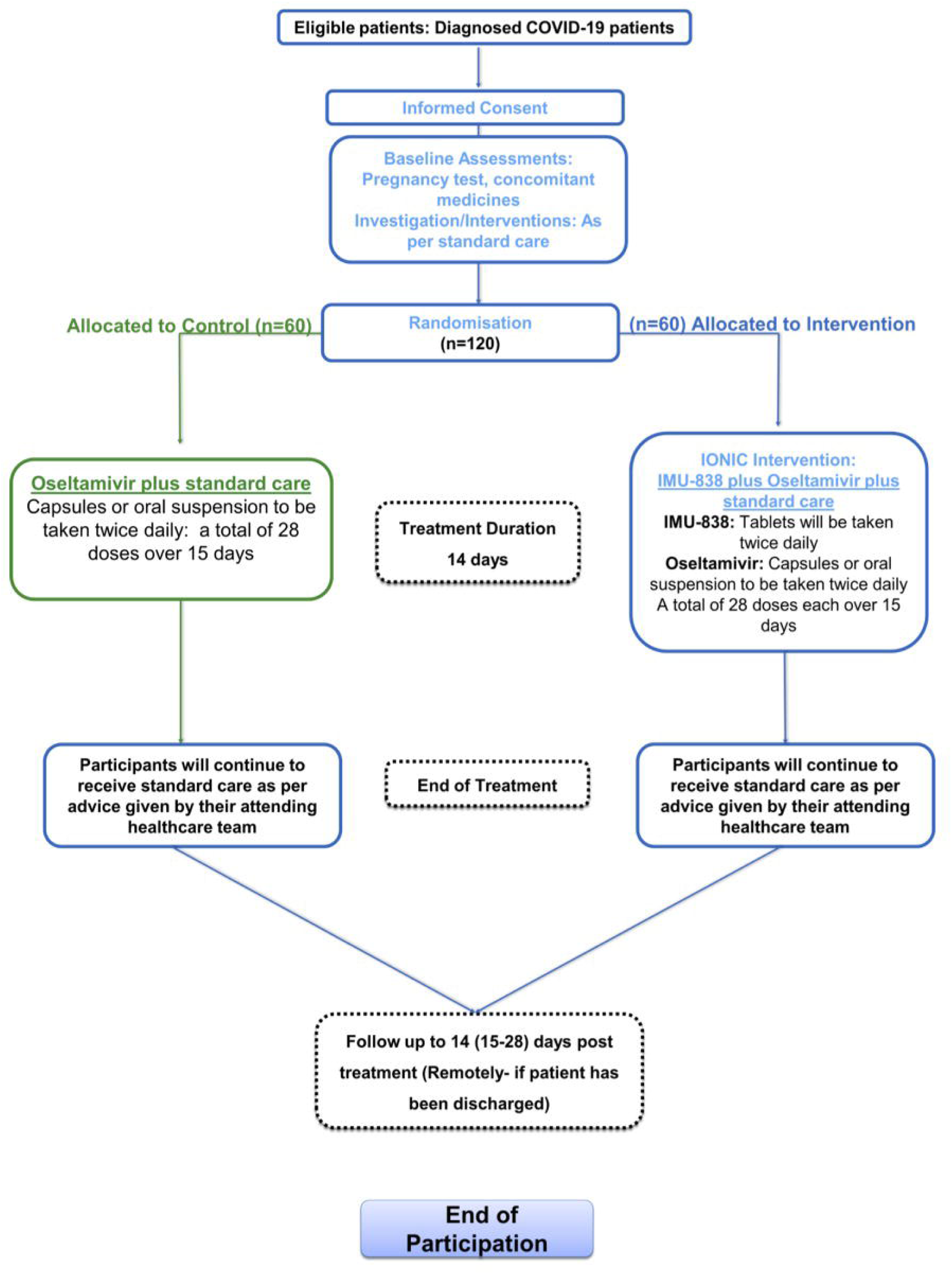
Flow of Participants in trial.

**Figure 3:**
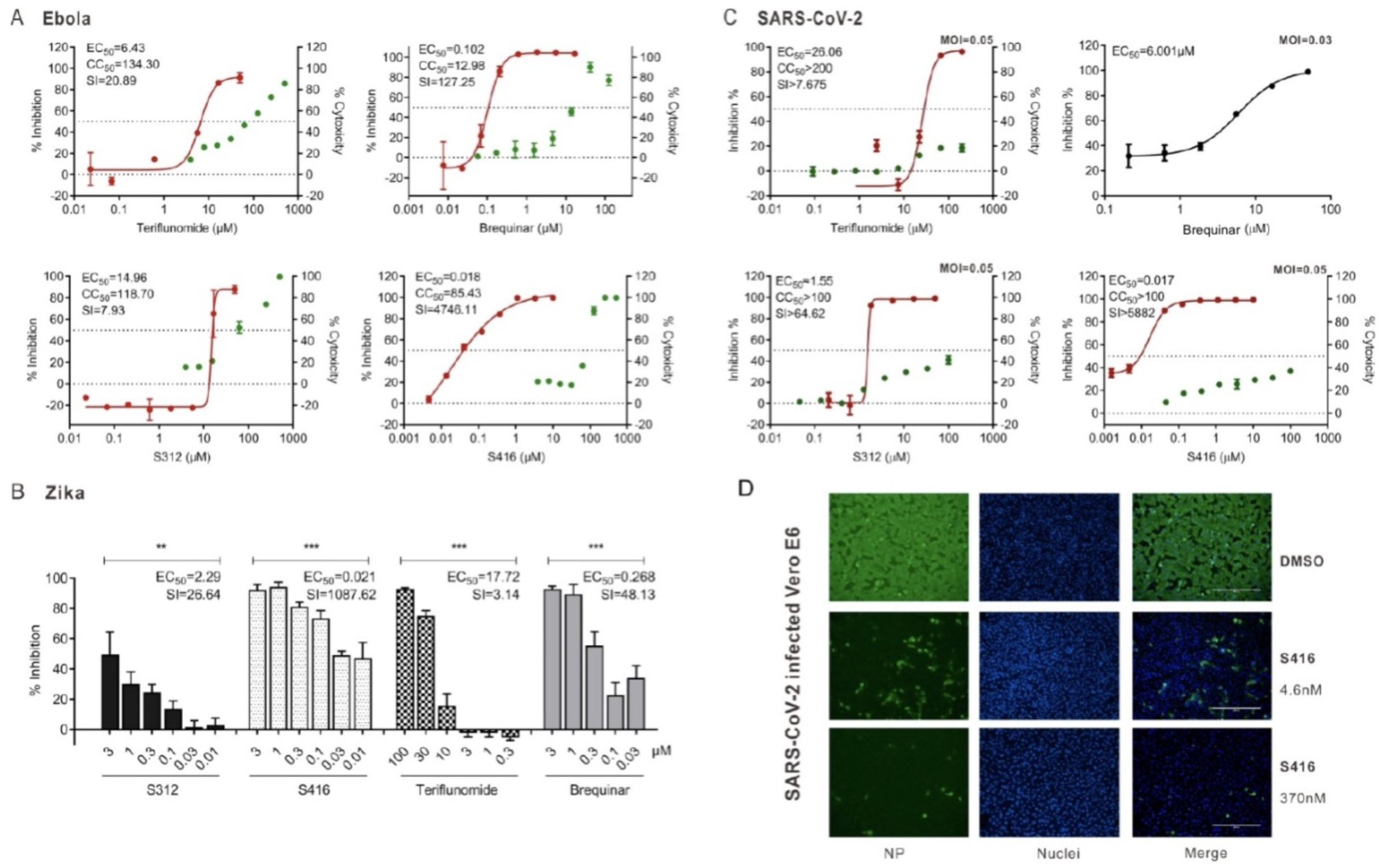
Antiviral activity of DHODH inhibitors (from Xiong et al. [2020]13) Note: (A) Anti-Ebola replication efficacy. BSR-T7/5 cells were transfected with the EBOV mini-genome replication system (NP, VP35, VP30, MG, and L) in the presence of increasing concentrations of Teriflunomide, Brequinar, S312 and S416 respectively. Inhibitory effects of these compounds (EC50) to EBOV mini-genome replication were determined using Bright-Glo Luciferase Assay (left-hand scale, red curve). CC50 of compounds were determined by analyzing BSR-T7/5 cell viability using CellTiterGlo Assay (righthand scale, green curve). The results are presented as a mean of at least two replicates ± SD. (B) Anti-Zika virus efficacy. Huh7 cells were infected with Zika virus (MOI=0.05) for 4 hours and then treated with increasing concentrations of compounds Teriflunomide, Brequinar, S312 and S416 respectively. The viral yields in cell supernatants were then quantified by qRT-PCR to reflect the replication efficiency of Zika virus. (C) Anti-SARS-CoV-2 virus efficacy. Aliquots of Vero E6 cells were seeded in 96-well plates and then infected with Beta CoV/Wuhan/WIV04/2019 at MOI of 0.03. At the same time, different concentrations of the compounds were added for co-culture. Cell supernatants were harvested 48 h.p.i. and RNA was extracted and quantified by qRT-PCR to determine the numbers of viral RNA copies. (D) Immunofluorescence assay of SARS-CoV-2-infected cells. Vero E6 cells were infected with SARS-CoV-2 under the same procedure of C. Cells were fixed and permeabilized for staining with anti-viral NP antibody, followed by staining with Alexa 488-labeled secondary antibody. Green represents infected cells. Nuclei were stained by DAPI, and the merge of NP and nuclei were shown. Scale bar, 400uM. The results (B, C) are presented as a mean of at least three replicates ± SD. Statistical analysis, One-way ANOVA for (B). NS, p >0.05; *, p <0.05; **, p <0.01; ***, p <0.001.

**Figure 4:**
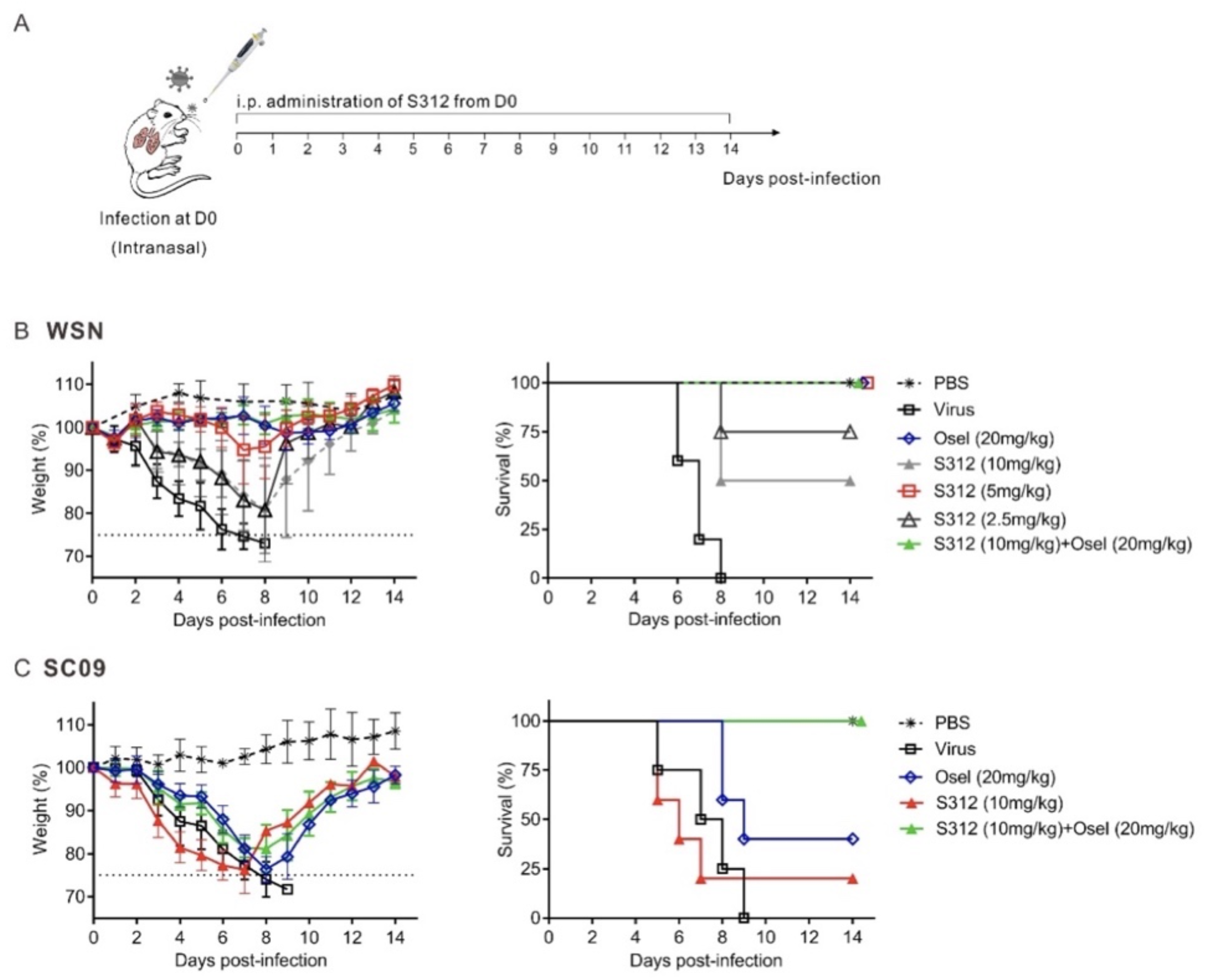
The in vivo antiviral activity of S312 in influenza A virus-infected mice (from Xiong et al. [2020]13) Note: (A) Diagram of the experimental procedure. (B) BALB/c mice were intranasal infected with 4000PFU of WSN virus and then intraperitoneal injected (i.p.) with PBS, S312 (2.5, 5, 10mg/kg), Oseltamivir (20mg/kg) and S312+Oseltamivir (10mg/kg+20mg/kg) once per day from D1-D14 respectively. The body weight and survival were monitored for 14 days or until body weight reduced to 75% (n = 4 mice per group). (C) Mice were inoculated intranasally with 600 PFU of A/SC/09 (H1N1) and then i.p. with S312 (10mg/kg), Oseltamivir (20mg/kg) and S312+Oseltamivir (10mg/kg+20mg/kg) once per day from D1 to D14. The body weight and survival were monitored until 14 days post-infection or when the bodyweight reduced to 75%. The dotted line indicates endpoint for mortality (75% of initial weight). The body weights are present as the mean percentage of the initial weight ±SD of 4-5 mice per group and survival curve were shown.

**Figure 5:**
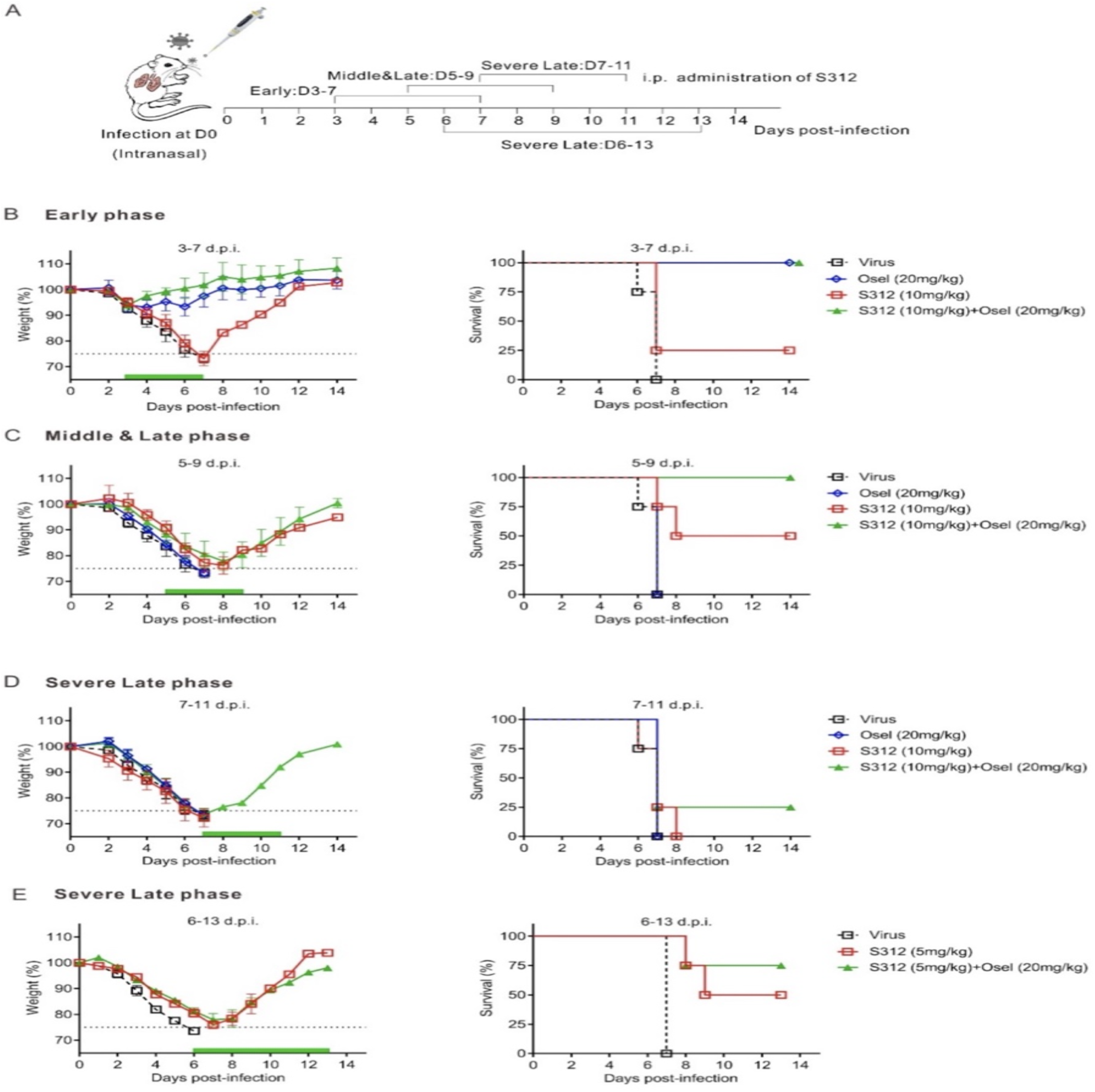
S312 is more effective at the late and severe infection phase as compared to the direct-acting antiviral drug oseltamivir (from Xiong et al. [2020]13) Note: (A) Diagram of the experimental procedure. (B-E) BALB/c mice were inoculated intranasally with 4000PFU of WSN virus and then i.p. with S312 (10mg/kg), Oseltamivir (20mg/kg), or S312+Oseltamivir (10mg/kg+20mg/kg) once per day from D3-7 (B), D5-9 (C), D7-11 (D). Another groups of S312 (5mg/kg) or S312+Oseltamivir (5mg/kg+20mg/kg) were given i.p. once per day from D6 to D13 in (E). The green bars indicate the period of drug administration. The body weight and survival were monitored until 14 days post-infection or when the bodyweight reduced to 75%, respectively (n = 4-5 mice per group). The dotted line indicates endpoint for mortality (75% of initial weight). The body weights are present as the mean percentage of the initial weight ± SD of 4-5 mice per group and survival curve were shown.

**Figure 6:**
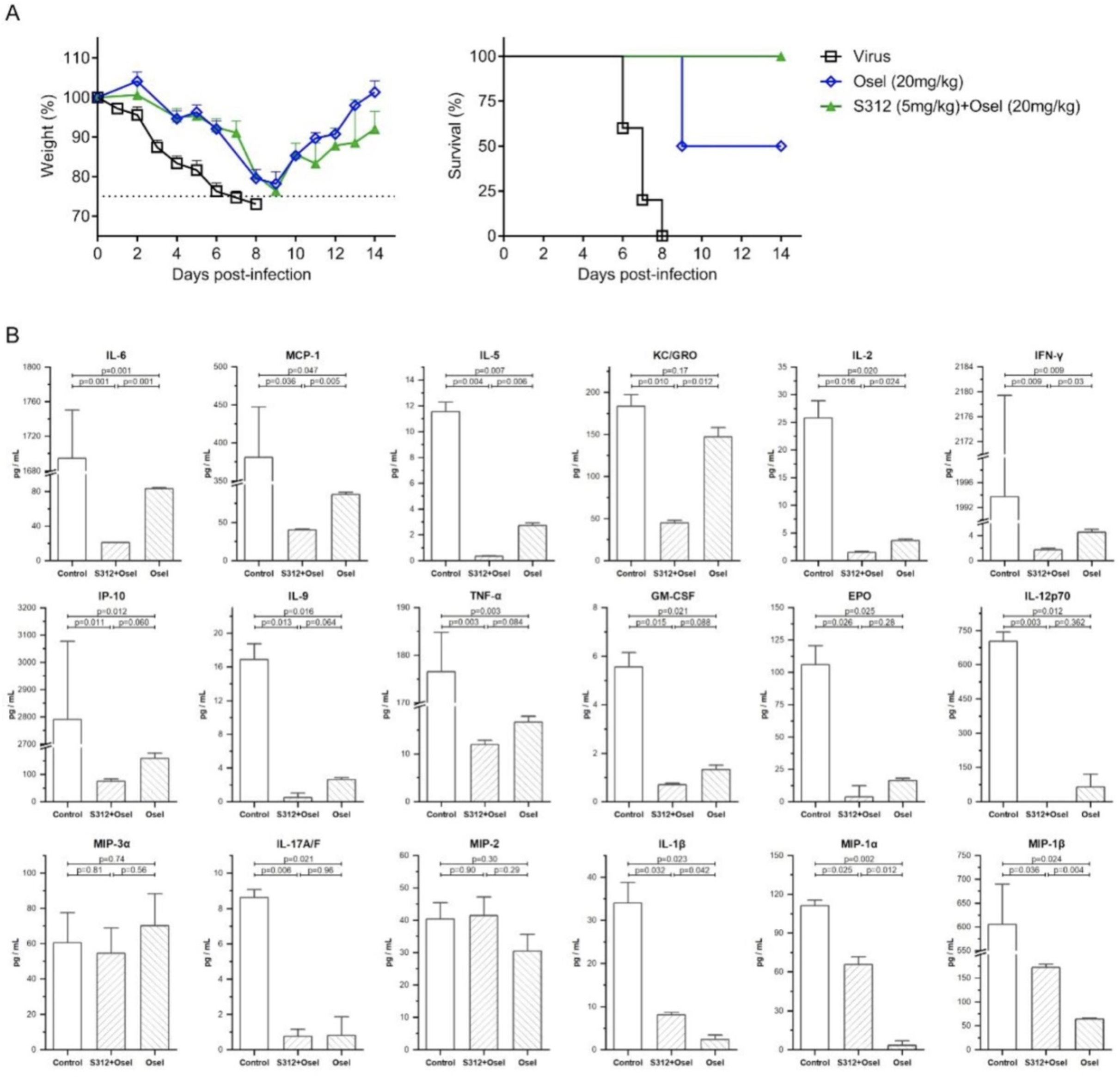
Cytokine and chemokine measurements following antiviral therapy (from Xiong et al. [2020]13) Note: (A) BALB/c mice were intranasally infected with 2000 PFU of influenza virus A/WSN/33 H1N1. Then, give mice intraperitoneal injection (i.p.) with Oseltamivir (20mg/kg), S312 + Oseltamivir (5mg/kg + 20mg/kg) once a day. Bodyweight loss and survival of the mice were monitored for 14 days or until body weight reduced to 75%, respectively (n = 5 mice per group). And dotted line indicates endpoint for mortality (75% of initial weight). (B) The cytokines and chemokines were measured by Meso Scale Discovery (MSD). The data were expressed as mean ± SD and were used to create the bar charts with error bars. The statistical analyses were performed using one-way ANOVA followed by Turkey post-hoc test. The plot function, ANOVA and the post-hoc functions were provided by OriginPro 2020 SR1 (9.7.0.188). P<0.05 was considered statistically significant and therefore the “significance level” parameters of the above functions were set to 0.05.

The IONIC trial comprises of a screening period, a 14-day treatment period, a 14-day follow-up period, and a long term follow up to one year evaluating the efficacy of IONIC intervention in comparison to Oseltamivir alone. All participants will receive standard care as necessary (e.g. supplemental oxygen, antibiotic agents vasopressor support etc.) in addition to IONIC Intervention or Oseltamivir, consistent with WHO recommendations. Treatment allocation will be assigned on a 1:1 ratio using variable block randomisation. After Day 14, all patients will continue with appropriate standard care as decided by the clinical care team.

The lead site of the study is University Hospital Coventry and Warwickshire NHS Trust. The study will be initiated as a single centre trial however, we are actively engaging with other NHS trusts which if interested will be invited to participate.

### 2.2. Screening and Consent

All patients admitted and hospitalised at UHCW with a confirmed or suspected case of COVID 19, that meet the eligibility criteria will be approached and offered the chance to participate in the IONIC trial.

Informed consent will be obtained from each patient before enrolment into the study. However, if the patient lacks capacity to give consent due to the severity of their medical condition then consent may be obtained from next of kin or friend acting as the patient’s personal legal representative. Further consent will then be sought with the patient if they recover sufficiently. Due to limitations on visitors on hospital premises consent will be taken verbally by telephone and documented on the consent form.

Due to the poor outcomes in COVID-19 patients who require ventilation (>90% mortality in one cohort ^(5)^), patients who lack capacity to consent due to severe disease (e.g. needs ventilation), and for whom a personal legal representative is not immediately available, randomisation and consequent treatment will proceed with consent provided by a treating clinician (independent of the clinician seeking to enrol the patient) who will act as the professional legal representative. Consent will then be obtained from the patient’s personal legal representative (or directly from the patient if they recover promptly) at the earliest opportunity.

### 2.3 Eligibility Criteria

**Inclusion criteria** will be any male or non-pregnant female who is 18 years or older with either: confirmed (positive result from a validated test) or suspected (has been in contact with a confirmed case of COVID 19 AND have mild to severe COVID 19 symptoms AND radiological evidence of pulmonary infiltrates) case of SARS-CoV-2. Hospitalisation must be in clinical status category 3-5 on the 9 point clinical status category scale proposed by WHO master protocol:

I. Category 3: hospitalized, no oxygen therapy
II. Category 4: hospitalized, oxygen by mask or nasal prongs
III. Category 5: hospitalized, non-invasive ventilation or high-flow oxygen

**Exclusion criteria** will be: anyone who is allergic or hypersensitive to IMU-838 or any of its ingredients; pregnant, breastfeeding or with the intention to become pregnant during the study, or participants who cannot take the trial medication orally at present. If the attending clinician specifies contraindication to the IONIC intervention or the patient has a specific medical or concomitant disease history preventing them to participate. In addition, if the participant is involved in any other interventional clinical trial for an experimental treatment of COVID 19.

### 2.4. Objectives and Outcome Measures/Endpoints

#### Primary objective

i. To evaluate the efficacy of IONIC Intervention (IMU-838 plus Oseltamivir and standard care) vs. Oseltamivir and standard care in adult participants with COVID-19 in relation to time-to-clinical improvement by 2 points on the 9 point WHO ordinal scale.

#### Secondary objectives

i. To evaluate safety and tolerability of *IONIC intervention* vs. Oseltamivir in adult subjects with COVID-19.
ii. To determine the effects of *IONIC Intervention* on improvement of at least two points in clinical status scale
iii. To assess the effects of IONIC Intervention vs. Oseltamivir on the need for invasive ventilation, renal replacement therapy or Extracorporeal membrane oxygenation (ECMO)
iv. To assess the effects of IONIC Intervention vs. Oseltamivir on the length of hospital and intensive care unit (ICU) stay
v. To assess the effects of (IONIC Intervention) vs. Oseltamivir on the time from treatment initiation to death.

#### Primary endpoints

i. Time-to-clinical improvement; defined as the time from randomisation to a 2-point improvement on WHO ordinal scale, discharge from hospital or death (whichever occurs first). Clinical status will be confirmed daily from randomisation to day 28, hospital discharge, or death (whichever occurs sooner), with the worst score for that day recorded.

#### Secondary endpoints

i. Adverse events (AEs) and serious adverse events (SAEs), including COVID-19 worsening and incidence of laboratory abnormalities
ii. Proportion of patients with two-point change on WHO ordinal scale at Day 7, 14 and 28 (± 2 days)
iii. Proportion of patients free of invasive ventilation, renal replacement therapy or ECMO at Day 7 and 14
iv. Hospital length of stay and Length of stay in Intensive care
v. Mortality at Day 28
vi. Time from treatment initiation to death (days)

### 2.5. Randomisation

Variable block randomisation will be carried out using an online validated randomisation sequence generator, as part of the Electronic Data Capture (EDC) system where the treatment allocation will be. The block sizes to be used in the randomisation sequence will be selected by the trial statistician.

Participants will be randomised on a 1:1 basis to IONIC Intervention or Control Group, stratified by Centre, Age groups and Sex. Data validation will be built into the EDC system to prevent randomisation unless the participant is eligible.

Only trained staff with the assigned user rights will be able to randomise participants using their unique username and password. An email notification will be automatically generated once the participant has been randomised. This email confirmation of the participant’s allocation will be sent to the Chief investigator and trial team.

Blinding and allocation concealment

This is an open-label study; therefore both the patients and trial staff will be aware of the patient’s allocated treatment. Allocation concealment will be maintained by using an independent online randomisation sequence generator.

### 2.6. Follow Ups

Follow-up information is to be collected on all study participants, irrespective of whether or not they complete the scheduled course of allocated study treatment. Study staff will seek follow-up information through various means (via telephone if discharged), including reviewing information from medical notes, routine healthcare systems, and registries.

Participants who are discharged during the course of treatment (14 days) and are continuing to take the Investigational medical product will be followed up remotely (via telephone) every 4 days (±24 hours) to monitor adverse events and drug compliance by a delegated research team member.

#### 2.6.1 Long Term Follow-up

There is emerging data to show that a percentage of patients experience long lasting effects of infection after recovering from COVID 19 infection referred to as ‘Long Covid’ ^21-22^. In an attempt to explore the prevalence of these long lasting effects in patients participating in the IONIC trial the study participants will be invited to remote follow-ups at 3 time points i.e. 3 months (±2 weeks), 6 months (±2 weeks) and 12 months (±2 weeks). Each follow up will record the participants WHO clinical status, health related quality of life questionnaire (EQ-5D-5L) and any further relevant medical history since discharge. All follow up activities will be conducted by a delegated member of the research team remotely and questionnaires will be delivered via telephone.

Prospective participants will have the option to only participate in the main trial by choosing not to participate in the long term follow up. A full schedule of events is available in table 1.

**Table 1:**
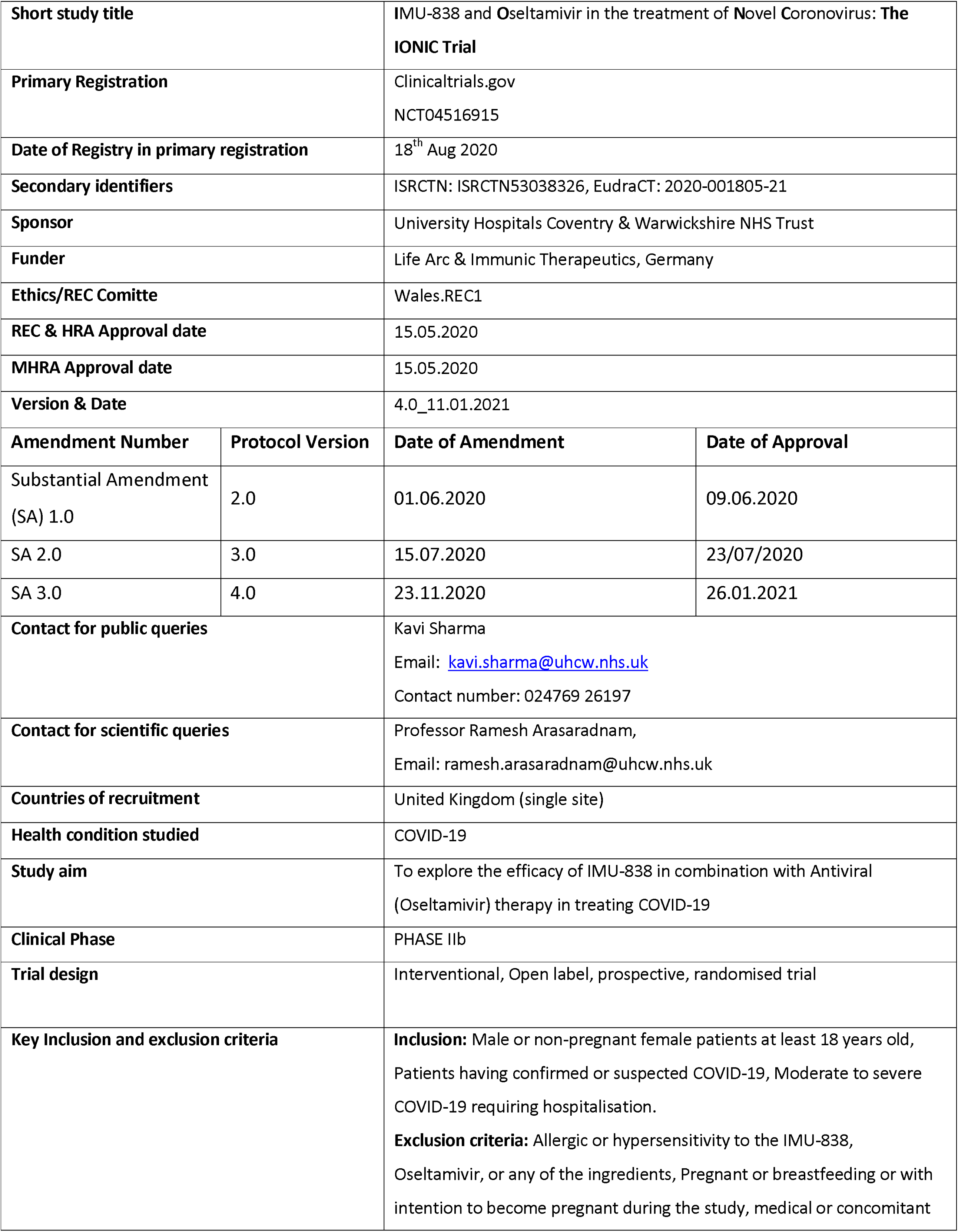
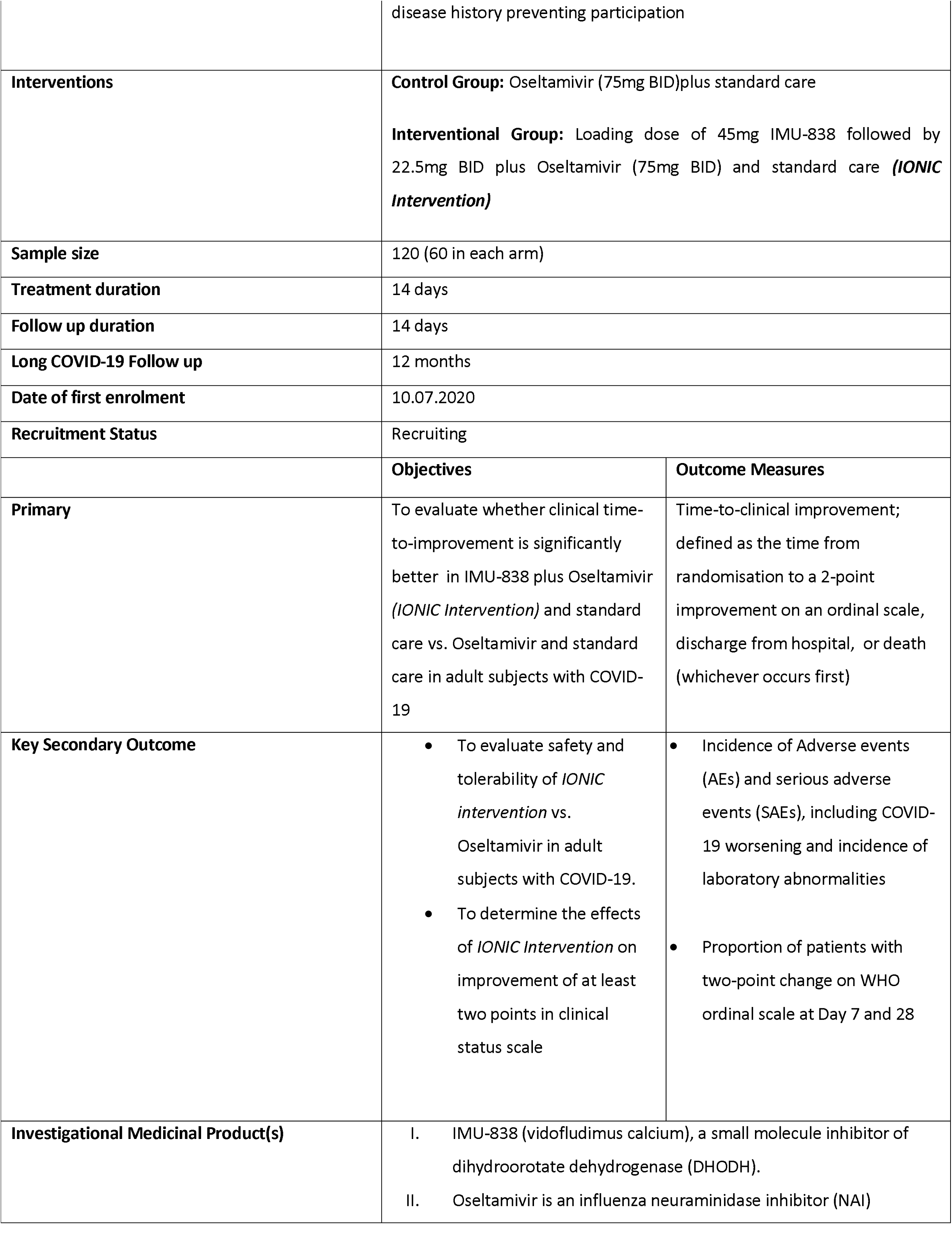
Administrative details and Trial Summary.

**Table 2:**
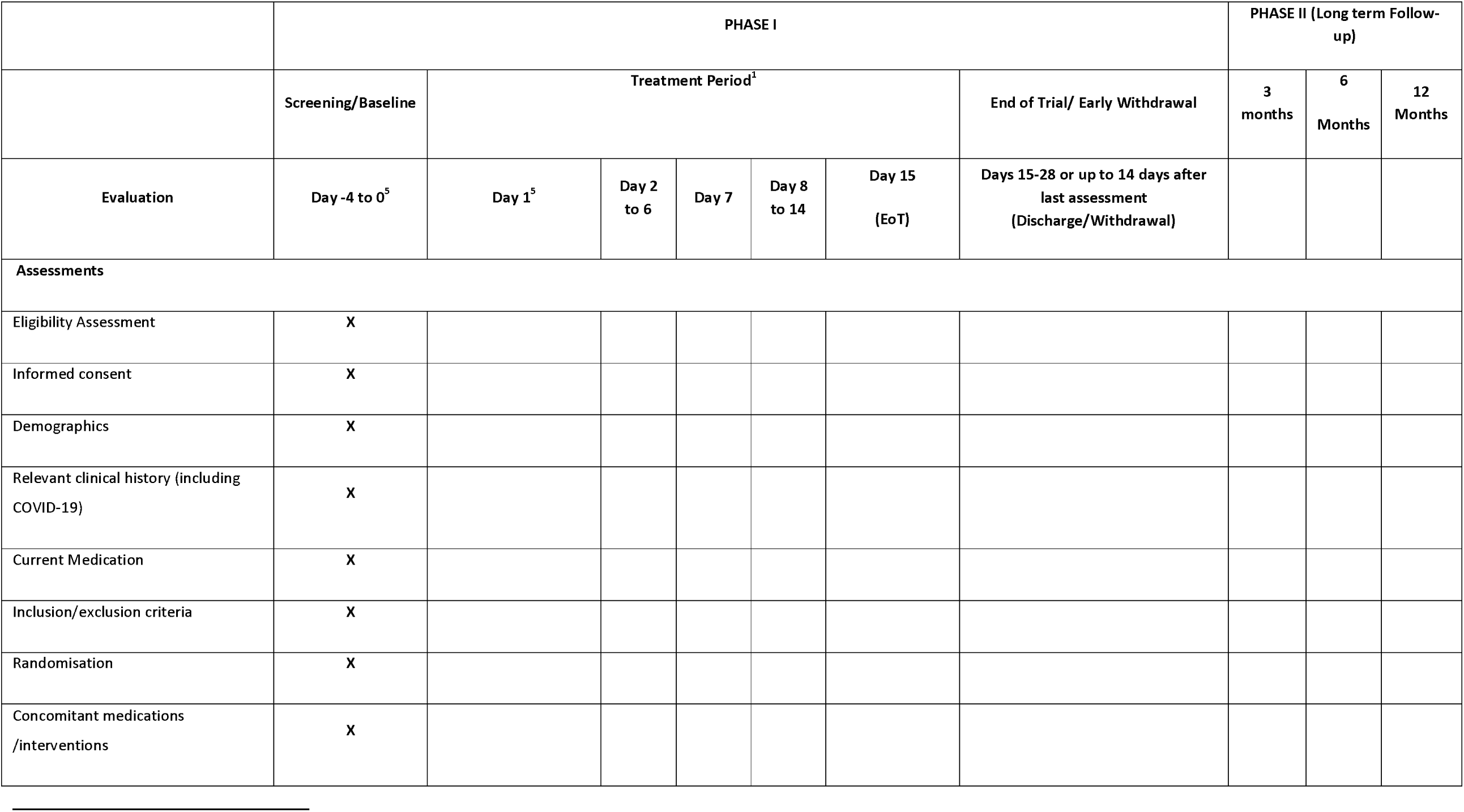
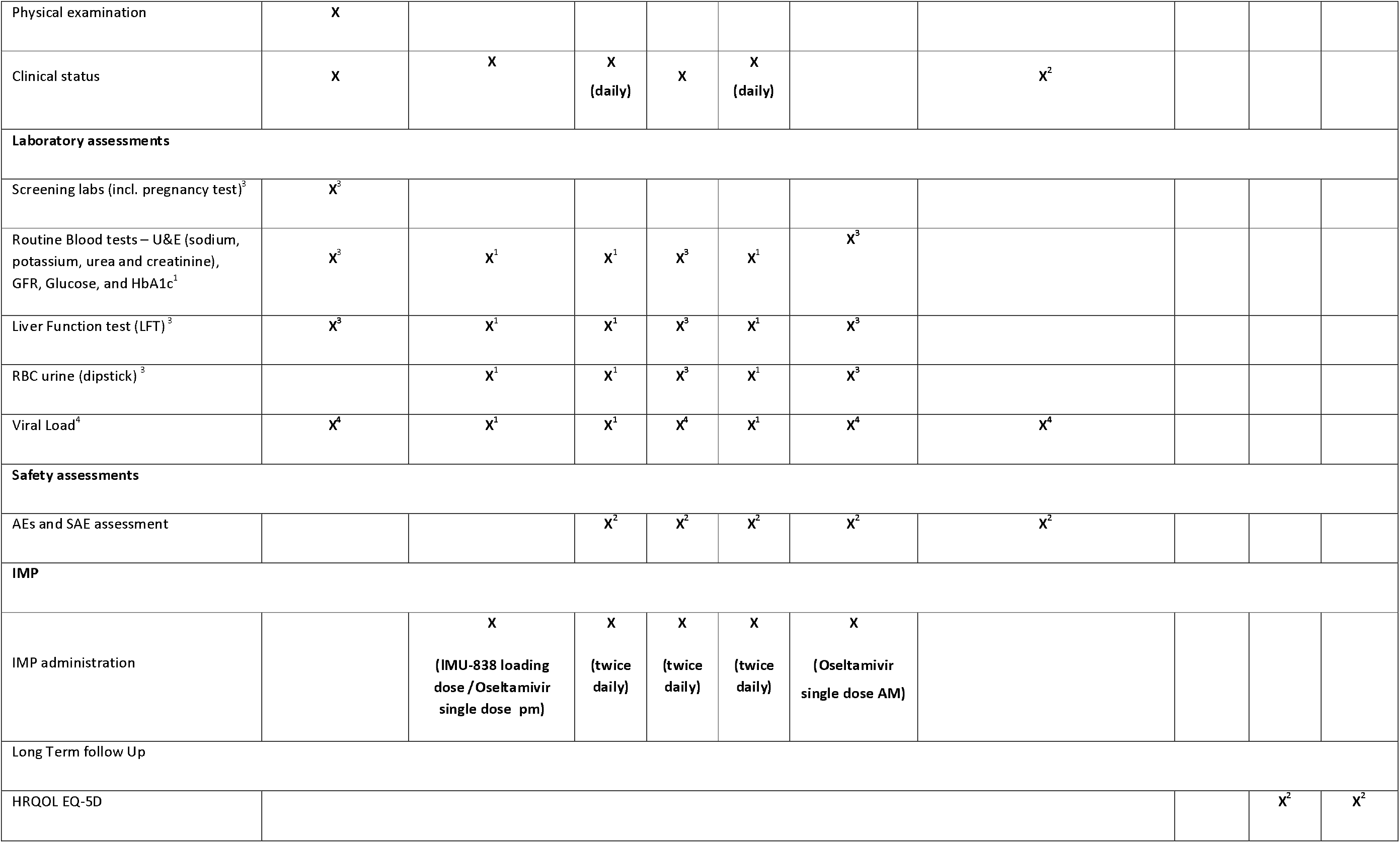
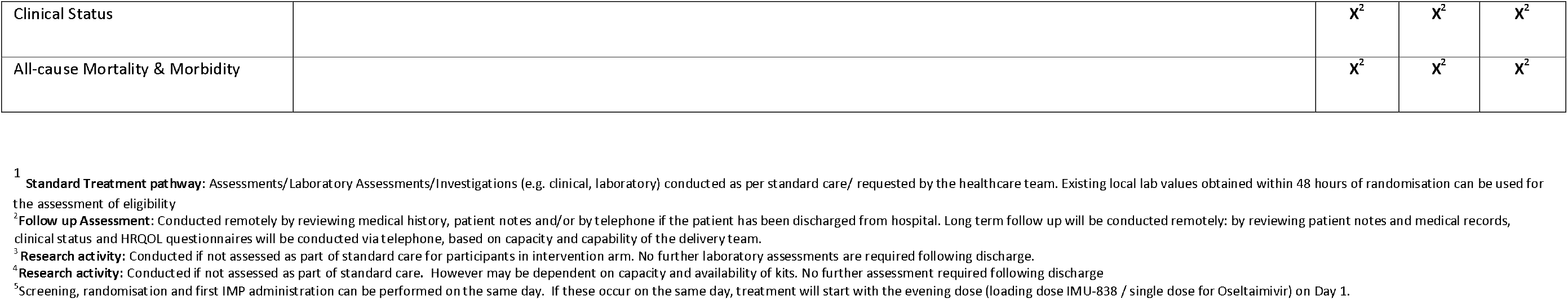
Schedule of events.

|  | PHASE I |  |  |  |  |  |  | PHASE II (Long term Follow-up) |  |  |
| --- | --- | --- | --- | --- | --- | --- | --- | --- | --- | --- |
|  | Screening/Baseline | Treatment Period<sup>1</sup> |  |  |  |  | End of Trial/ Early Withdrawal | 3 months | 6 Months | 12 Months |
| Evaluation | Day -4 to 0<sup>5</sup> | Day 1<sup>5</sup> | Day 2 to 6 | Day 7 | Day 8 to 14 | Day 15 (EoT) | Days 15-28 or up to 14 days after last assessment (Discharge/Withdrawal) |  |  |  |
| **Assessments** |  |  |  |  |  |  |  |  |  |  |
| Eligibility Assessment | X |  |  |  |  |  |  |  |  |  |
| Informed consent | X |  |  |  |  |  |  |  |  |  |
| Demographics | X |  |  |  |  |  |  |  |  |  |
| Relevant clinical history (including COVID-19) | X |  |  |  |  |  |  |  |  |  |
| Current Medication | X |  |  |  |  |  |  |  |  |  |
| Inclusion/exclusion criteria | X |  |  |  |  |  |  |  |  |  |
| Randomisation | X |  |  |  |  |  |  |  |  |  |
| Concomitant medications /interventions | X |  |  |  |  |  |  |  |  |  |
| Physical examination | X |  |  |  |  |  |  |  |  |  |
| Clinical status | X | X | X<br>(daily) | X | X<br>(daily) |  | X<sup>2</sup> |  |  |  |
| **Laboratory assessments** |  |  |  |  |  |  |  |  |  |  |
| Screening labs (incl. pregnancy test)<sup>3</sup> | X<sup>3</sup> |  |  |  |  |  |  |  |  |  |
| Routine Blood tests – U&E (sodium, potassium, urea and creatinine), GFR, Glucose, and HbA1c<sup>1</sup> | X<sup>3</sup> | X<sup>1</sup> | X<sup>1</sup> | X<sup>3</sup> | X<sup>1</sup> | X<sup>3</sup> |  |  |  |  |
| Liver Function test (LFT)<sup>3</sup> | X<sup>3</sup> | X<sup>1</sup> | X<sup>1</sup> | X<sup>3</sup> | X<sup>1</sup> | X<sup>3</sup> |  |  |  |  |
| RBC urine (dipstick)<sup>3</sup> |  | X<sup>1</sup> | X<sup>1</sup> | X<sup>3</sup> | X<sup>1</sup> | X<sup>3</sup> |  |  |  |  |
| Viral Load<sup>4</sup> | X<sup>4</sup> | X<sup>1</sup> | X<sup>1</sup> | X<sup>4</sup> | X<sup>1</sup> | X<sup>4</sup> | X<sup>4</sup> |  |  |  |
| **Safety assessments** |  |  |  |  |  |  |  |  |  |  |
| AEs and SAE assessment |  |  | X<sup>2</sup> | X<sup>2</sup> | X<sup>2</sup> | X<sup>2</sup> | X<sup>2</sup> |  |  |  |
| **IMP** |  |  |  |  |  |  |  |  |  |  |
| IMP administration |  | X<br>(IMU-838 loading dose /Oseltamivir single dose pm) | X<br>(twice daily) | X<br>(twice daily) | X<br>(twice daily) | X<br>(Oseltamivir single dose AM) |  |  |  |  |
| **Long Term follow Up** |  |  |  |  |  |  |  |  |  |  |
| HRQOL EQ-5D |  |  |  |  |  |  |  |  | X<sup>2</sup> | X<sup>2</sup> |

|  |  |  |  |  |
| --- | --- | --- | --- | --- |
| Clinical Status |  | X<sup>2</sup> | X<sup>2</sup> | X<sup>2</sup> |
| All-cause Mortality & Morbidity |  | X<sup>2</sup> | X<sup>2</sup> | X<sup>2</sup> |

<sup>1</sup> **Standard Treatment pathway:** Assessments/Laboratory Assessments/Investigations (e.g. clinical, laboratory) conducted as per standard care/ requested by the health care team. Existing local lab values obtained within 48 hours of randomisation can be used for the assessment of eligibility
<sup>2</sup> **Follow up Assessment:** Conducted remotely by reviewing medical history, patient notes and/or by telephone if the patient has been discharged from hospital. Long term follow up will be conducted remotely: by reviewing patient notes and medical records, clinical status and HRQOL questionnaires will be conducted via telephone, based on capacity and capability of the delivery team.
<sup>3</sup> **Research activity:** Conducted if not assessed as part of standard care for participants in intervention arm. No further laboratory assessments are required following discharge.
<sup>4</sup> **Research activity:** Conducted if not assessed as part of standard care. However may be dependent on capacity and availability of kits. No further assessment required following discharge
<sup>5</sup> Screening, randomisation and first IMP administration can be performed on the same day. If these occur on the same day, treatment will start with the evening dose (loading dose IMU-838 / single dose for Oseltamivir) on Day 1.

### 2.7. Patient withdrawal criteria

Patients must be withdrawn from the trial for any of the following reasons: Patient withdraws consent; investigator decision due to deterioration in renal or liver function (1.5 times increase in the values from baseline) which in the opinion of the investigator is not related to COVID-19; adverse event which, in the opinion of the investigator, may jeopardize the patient’s health or may compromise the trial objectives; relevant non-compliance with the protocol, which in the opinion of the investigator may jeopardize the trial integrity or scientific goals of the trial.

If the patient withdraws consent, no further evaluations should be performed and no attempts should be made to collect additional data. However, the patient may agree to continuing non-interventional follow-up procedures’.

Reasonable efforts will be made to contact any patient lost to follow up, to complete assessments and to retrieve any outstanding data and IMP and supplies. Patients who discontinue therapy with IMP will be encouraged to continue with trial-related assessments (including EoS visit) until their trial completion.

### 2.8 End of study definition

The end of the study will be defined at the date of the last participant’s End of Study assessment.

## 3. TRIAL TREATMENTS

### 3.1 IMU-838 (Vidofludimus calcium)

IMU 838 will be supplied by Immunic AG and will be manufactured, tested, and released according to current Good Manufacturing Practice guidelines and local requirements. IMU-838 will be administered twice daily as oral tablets starting with a loading dose of 45mg on the first day (Day 1, Table 3).

**Table 3:** Proposed dosing scheme for IMU-838 used in COVID-19 therapeutic trials.

|  | Day 1 |  | Day 2-13 |  | Day 14 |  |
| --- | --- | --- | --- | --- | --- | --- |
| Time | AM | PM | AM | PM | AM | PM |
| Number of Tablets |  | 2 | 1 | 1 | 1 | 1 |
| Dose of IMU-838 |  | 45 mg | 22.5 mg | 22.5 mg | 22.5 mg | 22.5 mg |
| | | $\alpha$ | $\beta$ | $\beta$ | $\beta$ | $\beta$ |

$\alpha$ Day1: loading dose of 45mg IMU-838 once daily given on the evening of Day 1
$\beta$ Day 2-14: dosing 22.g mg of IMU-838 BID

**Table 4.**
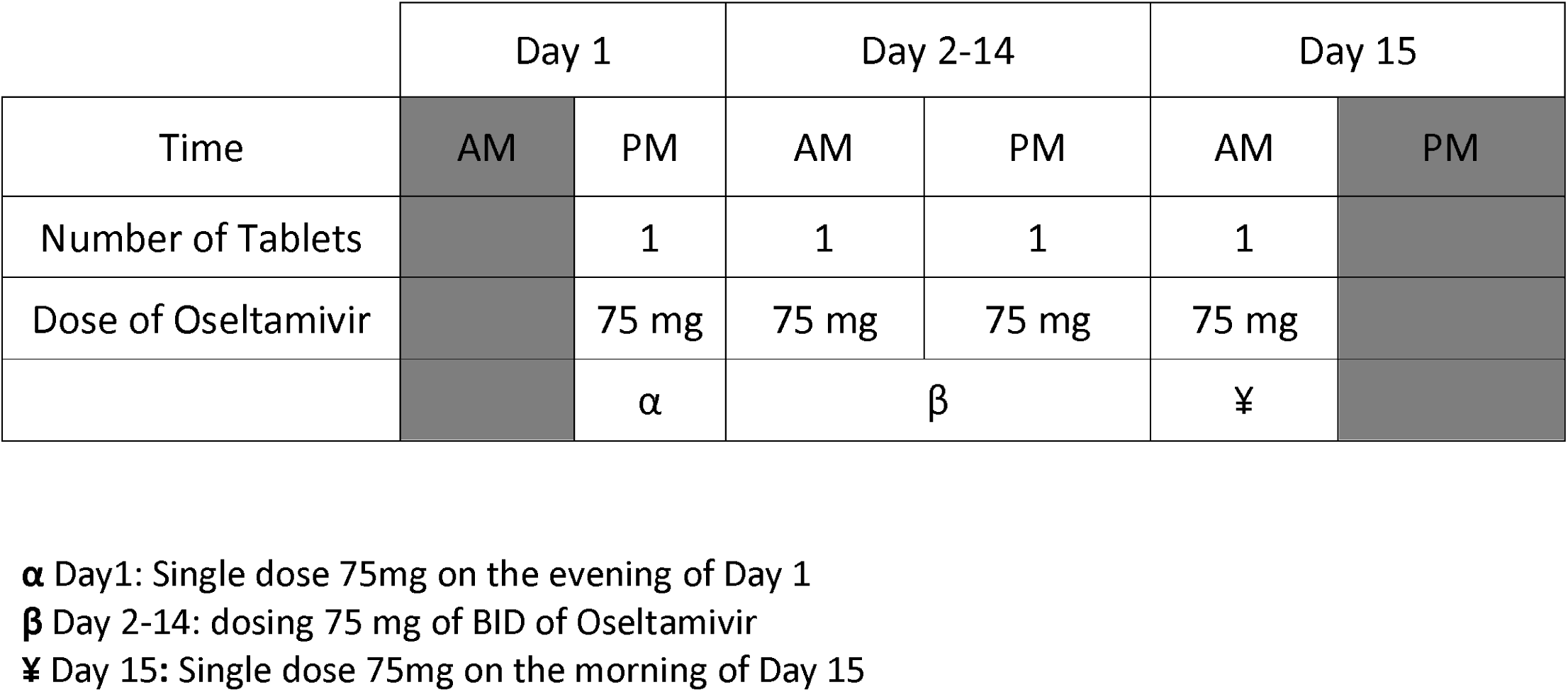
Proposed dosing scheme for Oseltamivir use in COVID-19 therapeutic trials.

Day 2-14: Once in the morning (15-60 min before a meal), and once in the evening (at least 2 hours after any meal and 15-60 min before any meal). The participants will be encouraged to drink sufficiently (approximately 1.5 litres per day throughout the trial).

### 3.2 Oseltamivir

Oseltamivir will be taken from commercially available stock with a UK Marketing Authorisation. 28 doses of Oseltamivir 75mg will be administered over 15 days as defined in the table below. Dose adjustments for renal impairment are outlined in Appendix 4.

## 4. STATISTICS AND DATA ANALYSIS

### 4.1 Sample size calculation

There will be a sample size of 60 participants in each arm of the study.

The sample size calculation is based on the analysis of the primary outcome, the time to clinical improvement. Current clinical knowledge suggests that patients in the control arm take about 14 days to improve by 2 points (i.e. a clinically significant improvement). Therefore, we motivated the power analysis using the expected percentage of the study population to have improved within the planned study follow-up time period of 14 days, under an assumed proportional hazards model.

We assume that 50% of patients in the control arm will improve within 14 days ^13^ and we hypothesise that 75% of patients will improve in the intervention arm; this results in a hazard ratio of 2 (see appendix). Using the standard formula for sample sizes of time-to-event outcomes ^13^ suggests that 52 patients are required in each arm of the study to detect a hazard ratio of this size with 80% power at the 5% level of significance. Allowing 10% loss to follow-up, the study would require approximately 120 participants.

### 4.2 Statistical analysis plan

The primary analysis will be on an intention-to-treat basis (i.e. as allocated), and will compare the time to clinical improvement between study arms using the proportional hazards survival model. The model will include terms to adjust for the status of the patients (ordinal assessment) at recruitment and other baseline data available such as age and sex. Patients who do not clinically improve or who die during the 14-days period will be right-censored. We will report hazard ratios and their 95% confidence intervals, and plot Kaplan-Meier curves to illustrate the time to improvement for both arms. For each intervention group and overall, we will report mean and standard deviation values (or proportions for dichotomous or ordinal measures) of baseline data. Analogous survival models will be fitted to the secondary outcomes that investigate time-to-event data. Linear regression models will be fitted to the continuous secondary outcomes. Secondary analyses will also include a per-protocol (i.e. as treated) analysis, and sensitivity analyses to explore the effect of the censored observations, due to death or deterioration, on the overall conclusions. All analyses will be undertaken in R4.0.0.

### 4.3 Interim Analysis

We will conduct key event analysis after data are available on 30 participants (15 in each arm) participants. This will allow the independent Data Monitoring Committee (DMEC) to make recommendations about adjustments to the study in the light of data on recruitment and outcome incidence, and to reassess our assumptions about sample size taking into account early data on the observed differences between the groups and safety information.

### 4.4 DATA MANAGEMENT

Trial data will be collected on CRFs and validated questionnaires, either on paper or electronically. An online validated, GCP compliant, Electronic Data Capture system will be used to record and store trial data. Individual user log-in access to this database will be granted to only those in the study team that require it for the performance of their role. Any paper copy of the CRFs and trial forms will be securely saved for 25 years in accordance with the UHCW NHS Trust archiving procedures. The information from these paper forms will also be recorded onto the database. All information stored on the database will be pseudonymised.

## Data Availability

Data from this study will be made available to researchers who provide a methodologically sound proposal in writing to the Sponsor, following the publication of the main study paper. Anonymised, individual participant data, data dictionary, study protocol and statistical analysis plan will be accessible upon application.

## 5. Declarations

### 5.1 DISSEMINATION POLICY

All data arising from the conduct of this study will remain the property of University Hospitals Coventry and Warwickshire NHS Trust. All efforts will be made to ensure that the results arising from the study are published in a timely fashion, in established peer-reviewed journals. Results will be disseminated to collaborators, colleagues, health professionals and participants via internal and external conferences and seminars, newsletters, and via interested groups, including local healthcare commissioning groups.

### 5.2 MONITORING, AUDIT & INSPECTION

The study will be monitored by the Research & Development Department at UHCW as representatives of the Sponsor, to ensure that the study is being conducted as per protocol, adhering to Research Governance and GCP. The approach to, and extent of, monitoring will be specified in a trial monitoring plan determined by the risk assessment undertaken prior to the start of the study.

### 5.3 Ethics approval

This study has been independently reviewed and approved by Wales Research Ethics Committee – (Ref No: 20/WA/0146); Health Research Authority (HRA) Approval was granted on 15/05/2020. In addition, required regulatory approvals were received from Medicines and Healthcare products Regulatory Agency (MHRA).

### 5.4 Consent for publication

Not applicable

### 5.6 Competing interests

The authors declare that they have no competing interests

### 5.7 Funding

The main phase of the study has received funding from LifeArc organisation. Immunic Therapeutics the manufacturer of IMU-838 has provided the funding for the trial drug used for this trial. The funding source had no role in the design of this study and will not have any role during its execution, analysis, interpretation of the data, or decision to submit results

### 5.8 Public and Patient Involvement

14 members of the UHCW Patient and Public Involvement (PPI) group reviewed the draft lay summary for this study, commenting on the concept of the study. The majority of reviewers confirmed that they would be ‘happy’ to take part or ‘had no objections’ to taking part in this study. The feedback was instrumental in designing the trial and producing the protocol.

A member of the UHCW PPI group was co-applicant on the funding application and continues to be part of the research team as a co-investigator, reviewing the trial design, protocol and additional documentation, and also being a member of the Trial Steering Committee.

All patients facing documentation has also been reviewed by members of the UHCW PPI group and feedback from this group has been taken into account in developing these documents.

### 5.9 Author contributions

AA and KS conceived of the presented idea. AA, LB and EV helped in developing the theory and delivery of the idea. NP and AN verified the analytical methods and the data analysis plan. LB and BL encouraged and assisted RA to investigate specific aspects [viral load] of the trial. KS has lead on the project management with significant support from BH and CB. TM has lead the research delivery team and assisted in recruitment. All authors discussed the results and contributed to the final manuscript.

## 5.10 Acknowledgements

Mr John Todd^2^

Dr Neerja Bhala^3,^

Dr Ravi Gowda, Prof Luca Frullon, Dr Mounia Hocine^4^

National Institute of Health Research (NIHR) Coventry and Warwickshire Clinical Research Facility^5^

The clinical research delivery team

Research participants

## Appendix 1

**Table 1:** Broad-spectrum antiviral efficacies of DHODH inhibitors (from Xiong et al. [2020](8)

| IC<sub>50</sub> (μM)<br>(SI*) | Virus type |  |  |  |  |
| --- | --- | --- | --- | --- | --- |
|  | H1N1<sup>a</sup> | H3N2<sup>b</sup> | H9N2<sup>c</sup> | Zika virus | Ebola-replicon |
| S312 | 2.36 (25) | 8.43 (7) | 13.2 (4) | 2.29 (26) | 15.0 (7.9) |
| S416 | 0.0161 (27) | 0.013 (126) | 0.020 (82) | 0.021<br>(1090) | 0.018<br>(4750) |
| Teriflunomide | 29.3 (6) | 2.73 (32) | 3.36 (26) | 17.7 (3) | 6.43 (32) |
| Leflunomide | >25.0 (<2.7) | NT | NT | NT | NT |
| Oseltamivir | 7.68 (680) | NT | NT | NT | NT |
| Brequinar | 0.240 (11.9) | 0.022 (130) | 0.060 (48) | 0.268 (48) | 0.102 (127) |

SI\* value was equal to CC<sub>50</sub>/IC<sub>50</sub>; values were rounded for significant figures from Xiong et al. 2020(8)
<sup>a</sup>A/WSN/33; <sup>b</sup>A/DongHu/06; <sup>c</sup>A/GuangZhou/99 NT (not tested)

## APPENDIX 2

### WHO Ordinal Scale for Clinical Improvement

#### Ordinal Scale for Clinical Improvement

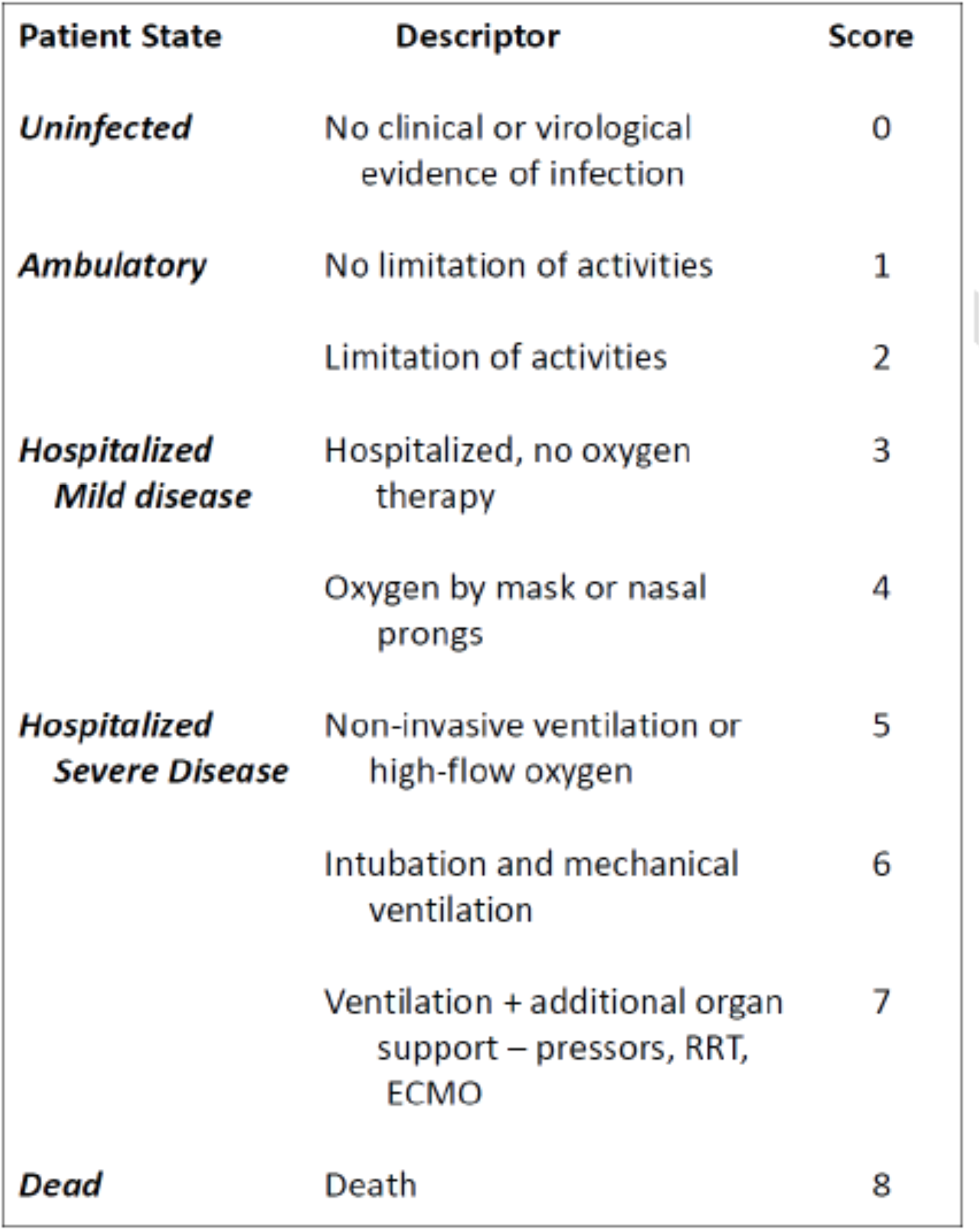

## Appendix 3

### Concomitant Medications & Medical History

#### 1. Therapy exclusion criteria

- Undergoing active chemotherapy or radiotherapy.
- Use of the following concomitant medications is prohibited at Screening Visit and throughout the duration of the trial:
  a. Use of Oseltamivir for more than 48 hrs prior to the first treatment dose
  b. Use of antiviral drugs (e.g. nucleoside analogue reverse-transcriptase inhibitors, protease inhibitors, etc.)
  c. History of long-term or concurrent use of mycophenolate mofetil, methotrexate exceeding 17.5 mg weekly
  d. Chloroquine or hydroxychloroquine
  e. Any medication known to significantly increase urinary elimination of uric acid, in particular lesinurad as well as uricosuric drugs such as probenecid
  f. Treatments for any malignancy, in particular irinotecan, paclitaxel, tretinoin, bosutinib, sorafinib, enasidenib, erlotinib, regorafenib, pazopanib and nilotinib
  g. Any drug significantly restricting water diuresis, in particular vasopressin and vasopressin analogues
  h. Use of rosuvastatin at daily doses higher than 10 mg

#### 2. Medical history and concomitant disease exclusion criteria

- Critical patients whose expected survival time < 48-72 hours
- Evidence of pancytopenia or immunosuppression
- Any contraindication to Oseltamivir or standard of care

Presence of the following laboratory values at Screening (samples taken to taken at Screening or any routine assessment performed within the last 5 days can be used to determine eligibility, where several the most recent should be reviewed):

- Platelet count <100,000/mm^3^ (<100 x 109/L)
  - Total bilirubin > 2 x ULN or ALT or GGT > 5 x ULN
  - Elevated indirect (unconjugated) bilirubin >1.2 x ULN (i.e. >1.1 mg/dL)
  - Serum uric acid levels at Screening Visit >1.2 x ULN (for women >6.8 mg/dL, for men >8.4 mg/dL)
  - Renal impairment defined as estimated glomerular filtration rate ≤30 mL/min/1.73m^2^
  - Decompensated liver cirrhosis (Child-Pugh score B and C)
  - History or presence of serious or acute heart disease such as uncontrolled cardiac dysrhythmia or arrhythmia, uncontrolled angina pectoris, cardiomyopathy, or uncontrolled congestive heart failure (New York Heart Association [NYHA] class 3 or 4) Note: NYHA class 3:
  - Cardiac disease resulting in marked limitation of physical activity. Patients are comfortable at rest. Less than ordinary activity causes fatigue, palpitation, dyspnea, or anginal pain. NYHA class 4: Cardiac disease resulting in inability to carry on any physical activity without discomfort. Symptoms of heart failure or the anginal syndrome may be present even at rest. If any physical activity is undertaken, discomfort is increased.
  - History or presence of any major medical or psychiatric illness (such as severe depression, psychosis, bipolar disorder), history of suicide attempt, or current suicidal ideation, if any of those conditions in the opinion of the investigator could create undue risk to the patient or could affect adherence with the trial protocol

#### 3. Women of child-bearing potential

If of child-bearing potential, must have a negative pregnancy test at Screening (blood test). They must agree not to attempt to become pregnant, must not donate ova, and must use a highly effective contraceptive method (see below) together with a barrier method between trial consent and 30 days after the last intake of the of investigational medicial product (IMP).

a. Highly effective forms of birth control are those with a failure rate less than 1% per year and include:
b. Oral, intravaginal, or transdermal combined (oestrogen and progestrogen containing) hormonal contraceptives associated with inhibition of ovulation
c. Oral, injectable, or implantable progestogen-only hormonal contraceptives associated with inhibition of ovulation
d. Intrauterine device or intrauterine hormone-releasing system
e. Bilateral tubal occlusion performed at least 6 months prior to study randomization
f. Vasectomised partner (i.e. the patient’s male partner underwent effective surgical sterilization before the female patient entered the clinical trial and is the sole sexual partner of the female patient during the clinical trial)
g. Sexual abstinence (acceptable only if it is the patient’s usual form of birth control/lifestyle choice; periodic abstinence [e.g. calendar, ovulation, symptothermal, postovulation methods] and withdrawal are not acceptable methods of contraception)
h. Barrier methods of contraception include:
  - Condom (without spermicidal foam/gel/film/cream/suppository or fat- or oil-containing lubricants)
  - Occlusive cap (diaphragm or cervical/vault caps) with spermicidal gel/film/cream/suppository

#### 4. Male participants of child bearing age

Male patients must agree not to father a child or to donate sperm starting at Screening Visit, throughout the clinical trial and for 30 days after the last intake of IMP. Male patients must also:

a. Abstain from sexual intercourse with a female partner (acceptable only if it is the patient’s usual form of birth control/lifestyle choice), or
b. Use adequate barrier contraception during treatment with IMP and until at least 30 days after the last intake of IMP, and
c. If they have a female partner of childbearing potential, the partner should use a highly effective contraceptive method as outlined above
d. If they have a pregnant partner, they must use condoms while taking IMP to avoid exposure of the foetus to the IMP

#### 5. Drug-drug interactions for IMU-838

a. The exposure to drugs metabolized by CYP2C8 may be increased by concomitant IMU-838 treatment. Concomitant administration of these drugs (especially those metabolized by more than 70% by CYP2C8) must, thus, be carefully considered. If possible, dose and treatment duration should be restricted or alternative drugs should be used. These drugs include:
  - Metabolized for more than 70% by CYP2C8: amodiaquine (anti-malarial), dasabuvir (anti-viral), enzalutamide (anti-cancer), montekulast (anti-asthmatic) and pioglitazone and repaglinide (anti-diabetics).
  - Metabolized for less than 70% by CYP2C8: paclitaxel, chloroquine, loperamide, ibuprofen and possibly diclofenac. In turn, strong CYP2C8 inhibitors such as gemfibrozil, glitazones, quercetin and trimethoprim may increase plasma concentrations of vidofludimus.
b. Medications with a metabolism and elimination being mainly dependent on CYP2C8 and CYP2C9 (with few alternative ways of elimination) should be taken with caution and should be monitored carefully. Given the known hepatotoxic potential of ibuprofen, the use of ibuprofen should be carefully considered or, if possible, therapeutic alternatives should be used.
c. The induction potential of IMU-838 for CYP1A2 may not lead to clinically relevant drug-drug interactions, however, they cannot be fully excluded. Although clopidogrel activation is performed via CYP1A2, the contribution of CYP1A2 is relatively small. It is known that some antipsychotic drugs, in particular clozapine, are partially eliminated via CYP1A2 and an induction of this enzyme may potentially reduce their drug efficacy.
d. In-vitro assays have shown synergistic effects of vidofludimus with infliximab.
e. Recent or concurrent treatment with uricosuric drugs such as probenecid or lesinurad may result in an increased risk of renal AEs since these drugs also inhibit URAT-1 and are expected to further elevate uric acid excretion. Therefore, uricosuric drugs should not be administered in combination with IMU-838. If uratelowering therapy is required, e.g. for gout flare prophylaxis, patients should be using xanthine oxidase inhibitors (allopurinol, febuxostat) or uricases (pegloticase, rasburicase) and should be monitored closely for changes in serum uric acid levels and renal function.
f. Because it cannot be excluded that vidofludimus interacts with protein binding of drugs that are strongly bound to plasma proteins, the plasma concentration of these drugs could be increased by vidofludimus. Similarly, vidofludimus plasma levels could increase by concomitant treatment with such drugs.
g. Vidofludimus has been shown in *in-vitro* studies to be a potent inhibitor of the organic anion transporters OAT1 and OAT3, and may therefore reduce the excretion of some drugs also using these transport systems.
h. IMU-838 is a strong inhibitor of BCRP (IC50 = 0.02 µM). If drugs that heavily depend on the BCRP transport system for elimination are co-administered with vidofludimus, patients should be closely monitored for signs and symptoms of excessive exposure to these drugs and their dosing should be carefully considered. This is particularly true for statins, and their dose should be lowered to the lowest possible dose. Specifically, doses of rosuvastatin are not to exceed 10 mg daily.
i. MTX doses of 17.5 mg/week or higher may slightly lower trough levels of vidofludimus and should not be used concomitantly with IMU-838.
j. Patients with UGT1A1 enzyme underexpression are at greater risk for irinotecan-induced severe diarrhea or neutropenia. Because vidofludimus inhibits UGT1A1, caution should be used when using vidofludimus in a patient undergoing therapy with irinotecan.

Further details can be found in the Investigator Brochure section 6.2.4.

## Appendix 4

### Dose adjustment in renal impairment

Considering that COVID-19 patients can suffer multi-organ failure which may include renal impairment the dose regime can be modified as per recommendations provided in the Renal Drug Database as follows, unless clinically indicated otherwise at the discretion of the treating physician:

| GFR | Recommended dose for treatment |
| --- | --- |
| > 60 (ml/min) | 75 mg twice daily |
| > 30 to 60 (ml/min) | 75 mg twice daily |
| > 10 to 30 (ml/min) | 75 mg once daily |
| ≤ 10 (ml/min) | 75mg as single dose<sup>*</sup> |

Note: This is in line with the Renal Drug database which differs from that in the SmPC and Public Health England and Scotland (updated September 2017) and is based on clinical experience and the good tolerability of oseltamivir.
\* In the event that GFR goes below the stated level and the participant has already received one or more doses as part of ongoing treatment during the trial they will not receive any more doses.

#### 8.2.6 Deterioration and requirement for NG tube

As the disease develops an NG tube may be inserted. For patients on the combination treatment arm, the IMU-838 tablets cannot be crushed and will not be administered via the NG tube. The oseltamivir will, however, continue to be administered.

The oseltamivir capsules can be opened and its contents mixed with a little bit of water for administration via an NG tube. The mixture should be stirred and given entirely to the patient. The mixture must be swallowed immediately after its preparation. For more details refer to the Oseltamivir SmPC (end of section 6.6). If the patent is discharged from the hospital before Day 14, the patient will receive the IMP(s) and will take the remaining doses of IMP at home. They will be given a medication card detailing their remaining treatment and administration.

## Appendix 5

### Consent Form

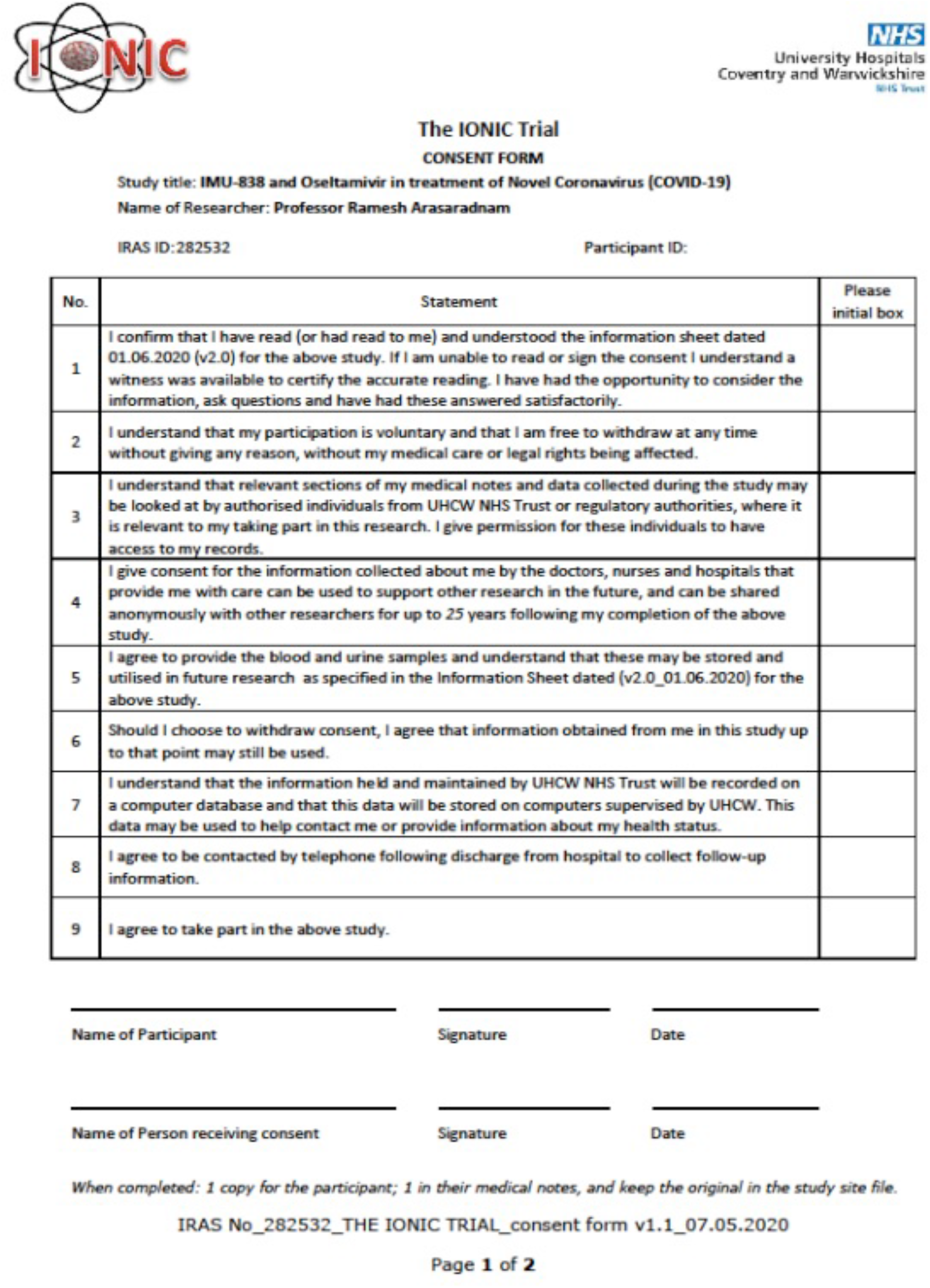

## Footnotes

2 Patient and public representative

3 Independent chair IONIC Data monitoring Committee / Trial Steering Committee

4 Independent Members IONIC Data monitoring Committee / Trial Steering Committee

5 This publication presents independent research funded by LifeArc and carried out with the support of the National Institute of Health Research (NIHR) Coventry and Warwickshire Clinical Research Facility. The views expressed are those of the author(s) and not necessarily those of LifeArc, the NHS, the NIHR or the Department of Health

## Notes

### Competing Interest Statement

The authors have declared no competing interest.

### Clinical Trial

NCT04516915
ISRCTN53038326

### Author Declarations

This study has been independently reviewed and approved by Wales Research Ethics Committee (Ref No: 20/WA/0146); Health Research Authority (HRA) Approval was granted on 15/05/2020. In addition, required regulatory approvals were received from Medicines and Healthcare products Regulatory Agency (MHRA).

